# The Impact of CYP2C19 Genotype on Clopidogrel Response and Clinical Outcomes in Coronary Artery Disease Patients: A Correlation with SYNTAX Score

**DOI:** 10.1101/2025.08.04.25332994

**Authors:** Sohaib Ashraf, Amjad Mujtaba, Amir Shahbaz, Shoaib Ashraf, Moneeb Ashraf, Rutaba Akmal, Muhammad Ahmad Imran, Sidra Ashraf, Meer Hassan, Qazi Abdul Saboor

## Abstract

**Background:** CYP2C19 polymorphism helps determine the reactivity of platelets in coronary artery disease. The impact of CYP2C19 genotype on clopidogrel response and clinical outcomes remains unclear. The SYNTAX score helps in determining the prognosis.

**Methods:** To test this, we enrolled more than 500 patients with ischemic heart disease. All the patients were on follow-up treatment of antiplatelets, clopidogrel and aspirin. We noted the CYP2C19 phenotype for cardiovascular events.

**Results:** This study included 156 individuals with a mean age of 59.2 ± 10.6 months and a median age of 60 months. Most patients were male (71.2%). The mean SYNTAX score was 26.1 ± 11.3, with a median of 29. Among participants, 25.6% had dyslipidemia, 75.6% had hypertension, and 49.4% had diabetes; only 9.6% had a history of PCI. There was no significant difference in age, BMI, sex, or smoking status between the stable angina and acute coronary syndrome groups. However, dyslipidemia, diabetes, PCI history, and a higher mean SYNTAX score were significantly more common in the acute coronary syndrome group. Statistical significance was achieved for SYNTAX score (P<0.05).

**Conclusion:** The SYNTAX score can be used to predict the complexity of CAD in patients. The greater the score, the greater the probability of CAD

## INTRODUCTION

Coronary artery disease (CAD) remains a leading global cause of death and is increasingly affecting younger individuals. Recent efforts focus on identifying and managing risk factors in young patients to lower CAD incidence and improve quality of life. While older patients with acute coronary syndrome often present with recognized risk factors, these are less commonly identified in younger patients with stable angina. (Sukhija et al., 2006). Coronary angiography is the gold standard for assessing atherosclerotic coronary artery disease but carries risks, ranging from minor complications to potentially life-threatening events if not promptly managed. Advances in procedural techniques, equipment design, and operator expertise have significantly reduced these risks over time. (Brilakis, 2021).

One in four Americans suffers from coronary heart disease. The American Heart Association reports that a heart attack occurs every 41 seconds in the United States. Coronary heart disease affects around 17 million Americans, with men comprising 55% of cases. Over half a million die from it annually in the U.S. Men by age 40 have a 49% lifetime risk, higher than women’s 32%. Although male predominance declines with age, coronary event incidence still rises as people get older. Globally, it remains the leading cause of adult death across all income levels (Sarkees & Bavry, 2009).

Normally angina symptoms include chest pain or an anginal analogue that is exacerbated by physical activity and relieved by rest or nitroglycerin. Even though there are several forms of angina, they are often characterized by symptoms like extreme fatigue, nausea, or shortness of breath in relation to the individual’s degree of activity (Boyette & Manna, 2022). The ACS and stable angina may require coronary arteriography or angiography to determine a better management plan.

Coronary arteriography has no absolute contraindications but carries risks of cardiac and non-cardiac complications. Certain patient factors—like severe CAD, heart failure with low ejection fraction, recent stroke or MI, and bleeding tendencies—can increase these risks. The likelihood of complications also depends on the type of procedure, whether diagnostic angiography alone or additional intervention (Brilakis, 2021). Serious complications from cardiac catheterization are rare, occurring in about 2% of patients, with a mortality risk below 0.08%. Advances like iso-osmolar contrast media, improved catheter design, better bleeding control, and skilled operators have further reduced these risks, allowing even critically ill patients to undergo the procedure safely when needed. (Manda & Baradhi, 2022).

A study conducted by Ramjattan in 2022 showed CT coronary angiography (CTCA) shows high accuracy, with 89% sensitivity and 96% specificity for detecting coronary heart disease. In early trials, 36% reported angina due to coronary disease, and 47% were diagnosed with CAD. CTCA is recommended for patients with chest discomfort and symptoms suggestive of coronary disease. (Ramjattan et al., 2022). The ESC/EACTS guidelines recommend the anatomic SYNTAX score (SxS) to assess coronary artery disease complexity. Initially developed from the SYNTAX trial to predict outcomes after PCI or CABG in stable patients, the SxS has since been applied to other patient groups, including those undergoing primary PCI for acute coronary syndromes. (Y.-H. Kim et al., 2010).

Studies, including SYNTAX trial sub-analyses, show that the original SYNTAX score often misclassifies risk, especially for mortality in stable CAD or ACS patients treated with PCI. To improve risk stratification, the SYNTAX score II (SxS II) was developed by adding clinical factors like age, creatinine clearance, LVEF, unprotected left main disease, peripheral vascular disease, gender, and COPD. This enhanced model offers better accuracy, particularly in patients with left main or multivessel disease. (Scherff et al., 2011).

Cardiologists, interventionists, and surgeons all utilize the SYNTAX score, which is based on an angiographic approach, to determine the degree of difficulty of coronary artery lesions. Higher scores predict poorer outcomes after modern revascularization, especially PCI. While it shows some predictive value for major adverse cardiac and cerebrovascular events, its role in forecasting early or late mortality remains unclear. Evidence supports its use in left main PCI, but data are limited for predicting outcomes in patients with three-vessel disease undergoing PCI or stenting. (Safarian et al., 2014a).

The rationale of this study is to compare the angiographic findings in stable angina versus acute coronary syndrome by using the SYNTAX score. The results of this study will benefit in the better management of these patients.

## MATERIAL AND METHODS

This prospective comparative study was conducted over 12 months at the Department of Cardiology, Shaikh Zayed Hospital, Lahore, and included 156 patients undergoing coronary angiography. Participants were divided into two groups: Group A (78 patients with stable angina) and Group B (78 patients with acute coronary syndrome). The sample size was calculated using proportions from a previous study published in 2019 (See Supplementary File) at 80% power and 95% confidence interval, accounting for a 10% dropout rate [Fukui, T, et al., 2013]. Patients aged 18–80 years of both genders undergoing primary PCI or elective coronary angiography were included. Patients with a history of prior cardiac catheterization, PCI, or CABG were excluded. The exclusion criteria also included a previous history of cardiac catheterization and PCI patients with CABG.

Approval was taken from the Institutional Review Board (IRB) of Shaikh Zayed Medical Complex, Lahore, total of 156 patients who fulfilled the inclusion criteria and were admitted through the OPD and the emergency Department of the Shaikh Zayed Hospital, Lahore. A written informed consent was taken from each patient after explaining the risks, purpose, and benefits of this research. Detailed history, including age, gender, height, weight, BMI, and medical number, was noted, and SYNTAX score was used after angiography for data collection for both groups. The selected patients were classified into two groups (A & B). Group A patients were referred as coronary angiography with stable angina, and other patients in group B was referred to acute coronary syndrome. The patients were instructed to follow up after 30 days of discharge if any complications develop after this clinical procedure.

Data was analyzed by using SPSS v.25. Mean ± SD were computed for quantitative variables like age, hypertension and BMI. Qualitative variables like gender, diabetic mellitus, PCI, were presented as frequency and percentages. Post-stratification, chi-square test was applied to compare the factors in stratified groups. P ≤0.05 was considered as significant.

## RESULTS

A total of 156 patients were included in the study. The majority of participants were male (n = 111, 71.2%), while females accounted for 28.8% (n = 45). Patients were equally distributed into two clinical groups: Group A with stable angina (n = 78, 50.0%) and Group B with acute coronary syndrome (ACS) (n = 78, 50.0%). Regarding smoking status, most participants were non-smokers (n = 128, 82.1%), whereas smokers comprised 17.9% (n = 28). Dyslipidemia was present in 25.6% (n = 40) of the study population, while the remaining 74.4% (n = 116) did not have dyslipidemia. The study included 118 (75.6%) hypertensive patients and (n=77. 49.4%) diabetes patients. However, only 15 (9.6%) patients underwent PCI (Table 1, Figure 1).

**Figure 1.**
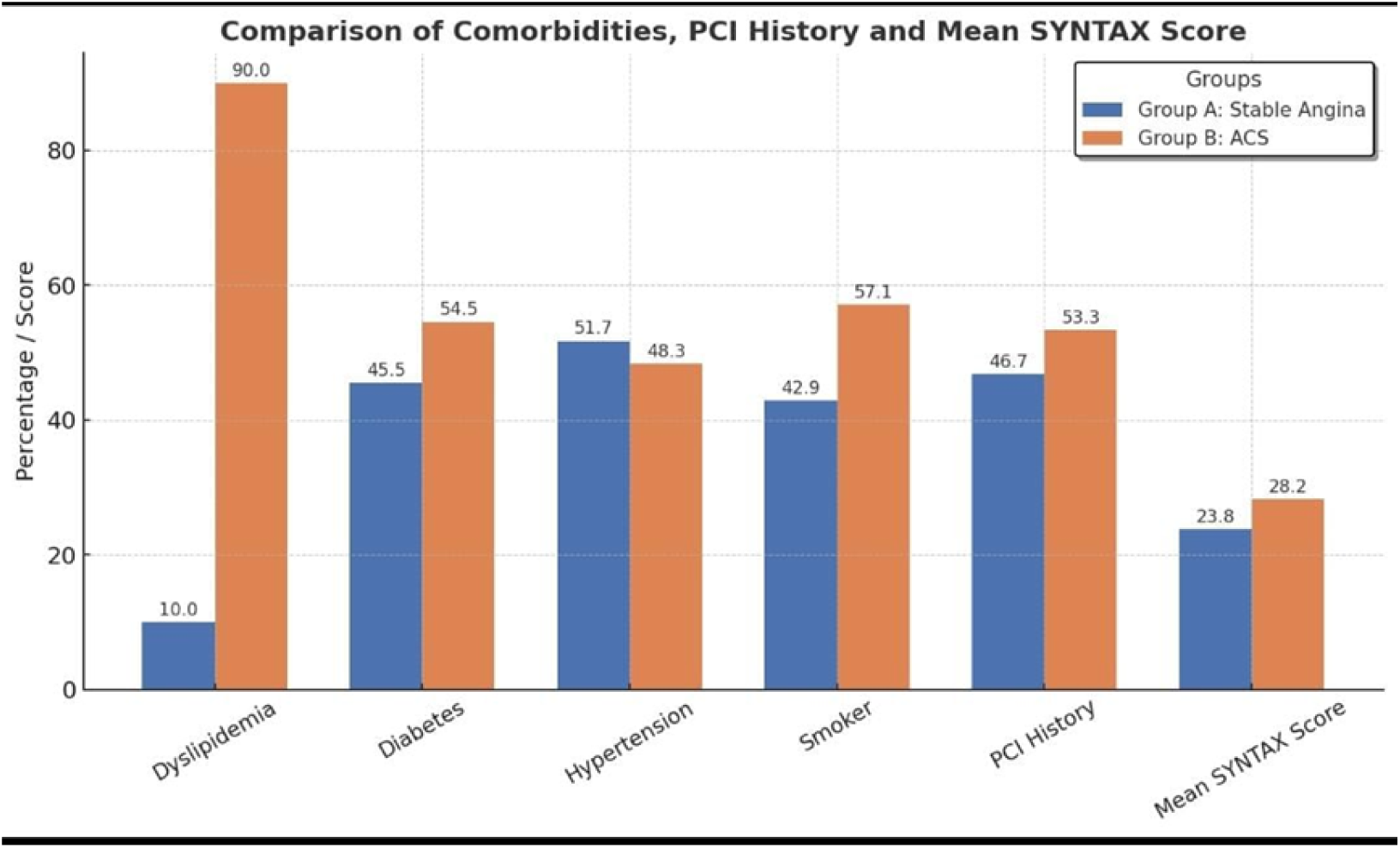
Comparison of comorbidities, prior PCI history, and mean SYNTAX scores between Group A (Stable Angina) and Group B (Acute Coronary Syndrome, ACS). The figure illustrates the prevalence of dyslipidemia, diabetes, hypertension, and smoking status, along with previous PCI history and the mean SYNTAX score in each group. Patients with ACS demonstrated a notably higher prevalence of dyslipidemia and smoking, as well as higher mean SYNTAX scores compared to those with stable angina

**Table 1.**
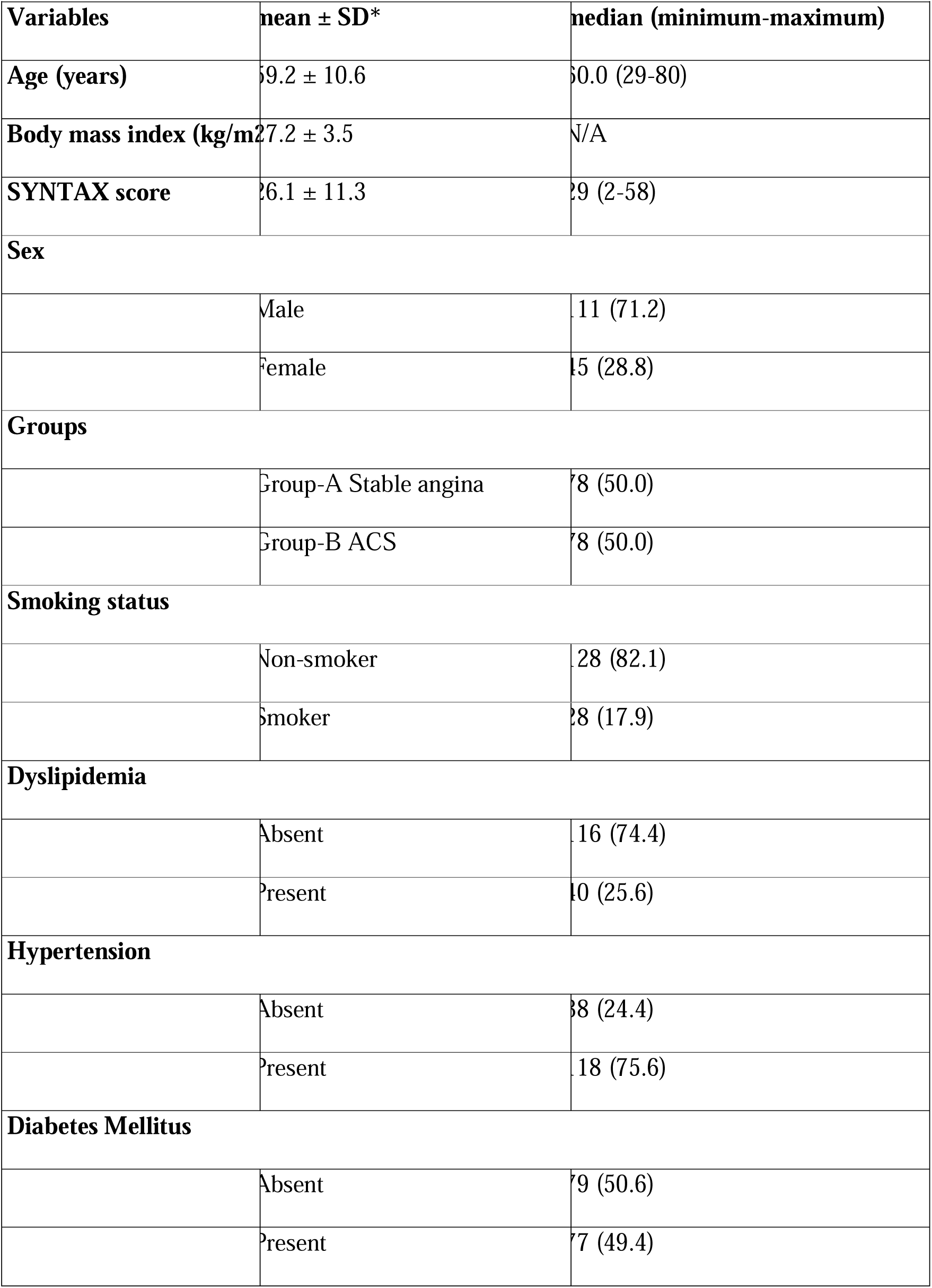
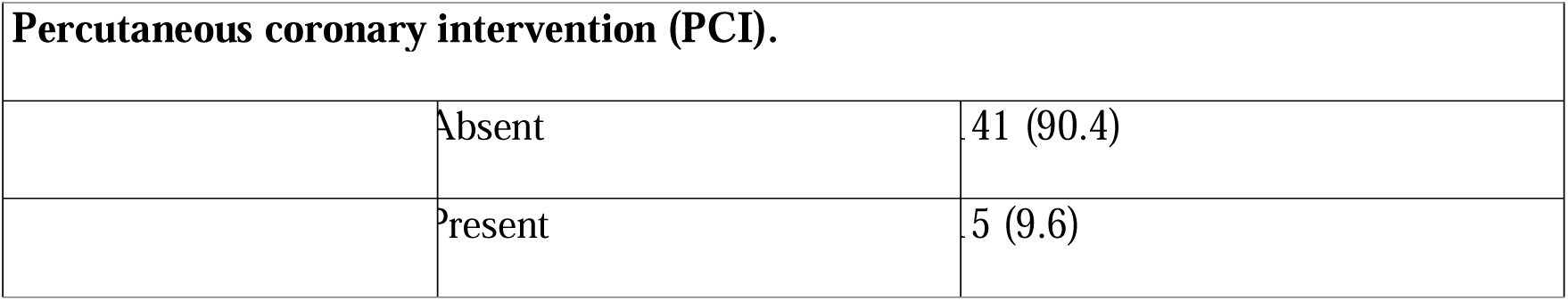
Baseline characteristics of study participants.

The mean age of the patients was 59.2 ± 10.6 years, with a median age of 60.0 years (range: 29–80 years). The mean body mass index (BMI) was 27.2 ± 3.5 kg/m². The angiographic assessment using the SYNTAX score revealed a mean score of 26.1 ± 11.3, with a median of 29 (range: 2–58). These findings describe the baseline demographic and clinical characteristics of the study cohort (Table 1, Figure 1).

The study cohort was equally divided into Group A (stable angina, n = 78) and Group B (acute coronary syndrome, n = 78). The mean age of patients in Group A was slightly higher (60.2 ± 10.1 years) compared to Group B (58.3 ± 11.0 years), although this difference was not statistically significant (p = 0.26). Similarly, the mean body mass index (BMI) was comparable between the two groups (27.1 ± 2.2 kg/m² in Group A vs. 27.4 ± 4.4 kg/m² in Group B; p = 0.58). However, the SYNTAX score was significantly higher in patients with acute coronary syndrome (mean 28.2 ± 9.9) than those with stable angina (mean 23.8 ± 12.4), with a p-value of 0.01, indicating greater angiographic complexity in Group B (Table 2).

**Table 2.**
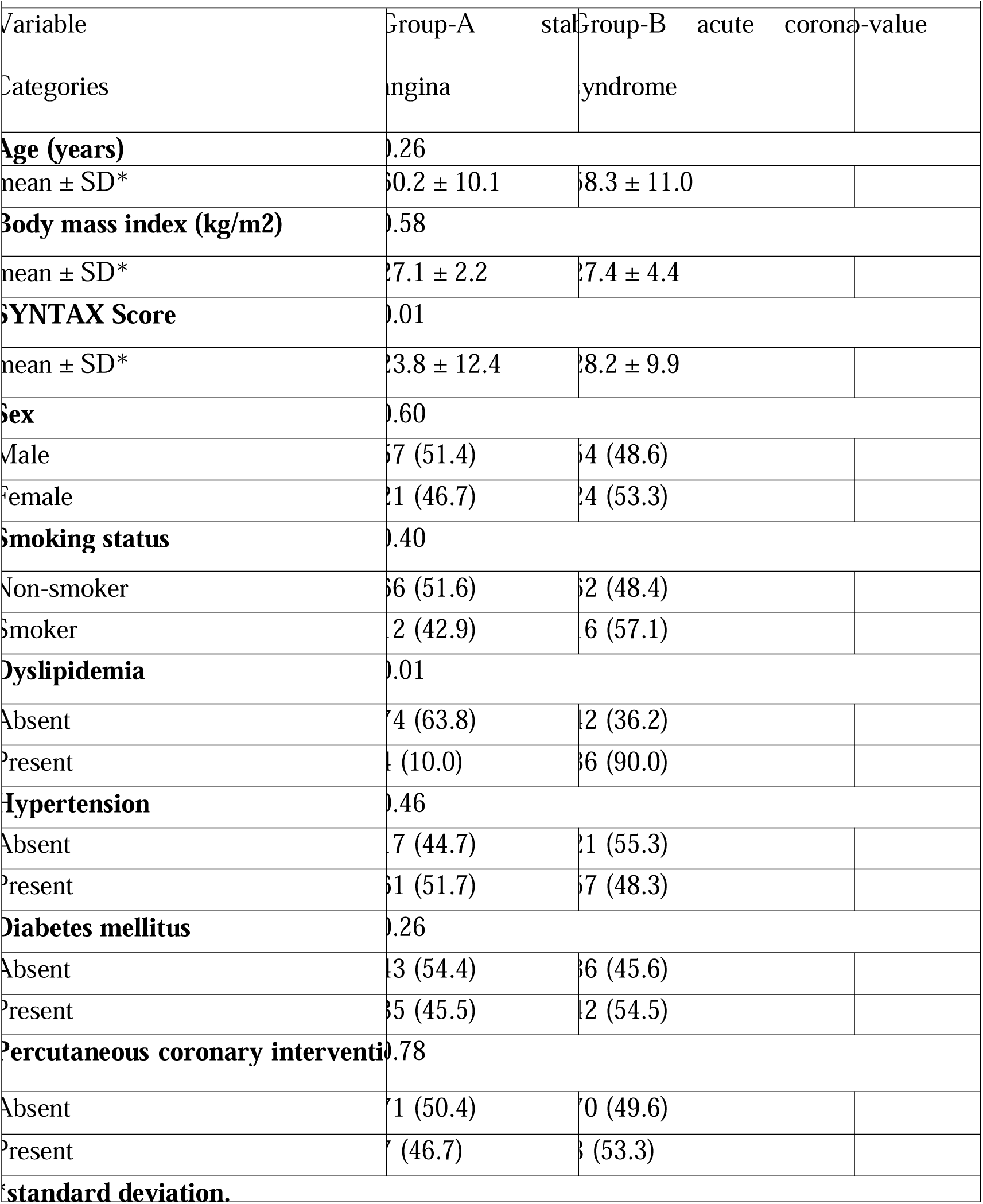
Bifurcation of demographic variables with respect to Group-A Stable angina versus Group-B acute coronary syndrome.

The sex distribution in males was predominated in both groups (51.4% in Group A vs. 48.6% in Group B; p = 0.60). The proportion of smokers was higher in Group B (57.1%) compared to Group A (42.9%), but this difference was not statistically significant (p = 0.40). This represents the homogeneity in the groups. The dyslipidemia in Group B had a greater prevalence compared to Group A, and the results were statistically significant. (90.0% 10.0%, P<0.05). Hypertension was common in both groups, present in 51.7% of Group A and 48.3% of Group B patients, with no significant difference (p = 0.46). Similarly, diabetes mellitus was more common among group B than group A, but the results were not statistically significant (54.5%, 45.5%, P>0.05). There was no significant difference in age, BMI, sex, or smoking status between the stable angina and acute coronary syndrome groups. However, dyslipidemia, diabetes, PCI history, and a higher mean SYNTAX score were significantly more common in the acute coronary syndrome group. And statistical significance was achieved for SYNTAX score (P<0.05) (Table 2, Figure 2).

**Figure 2.**
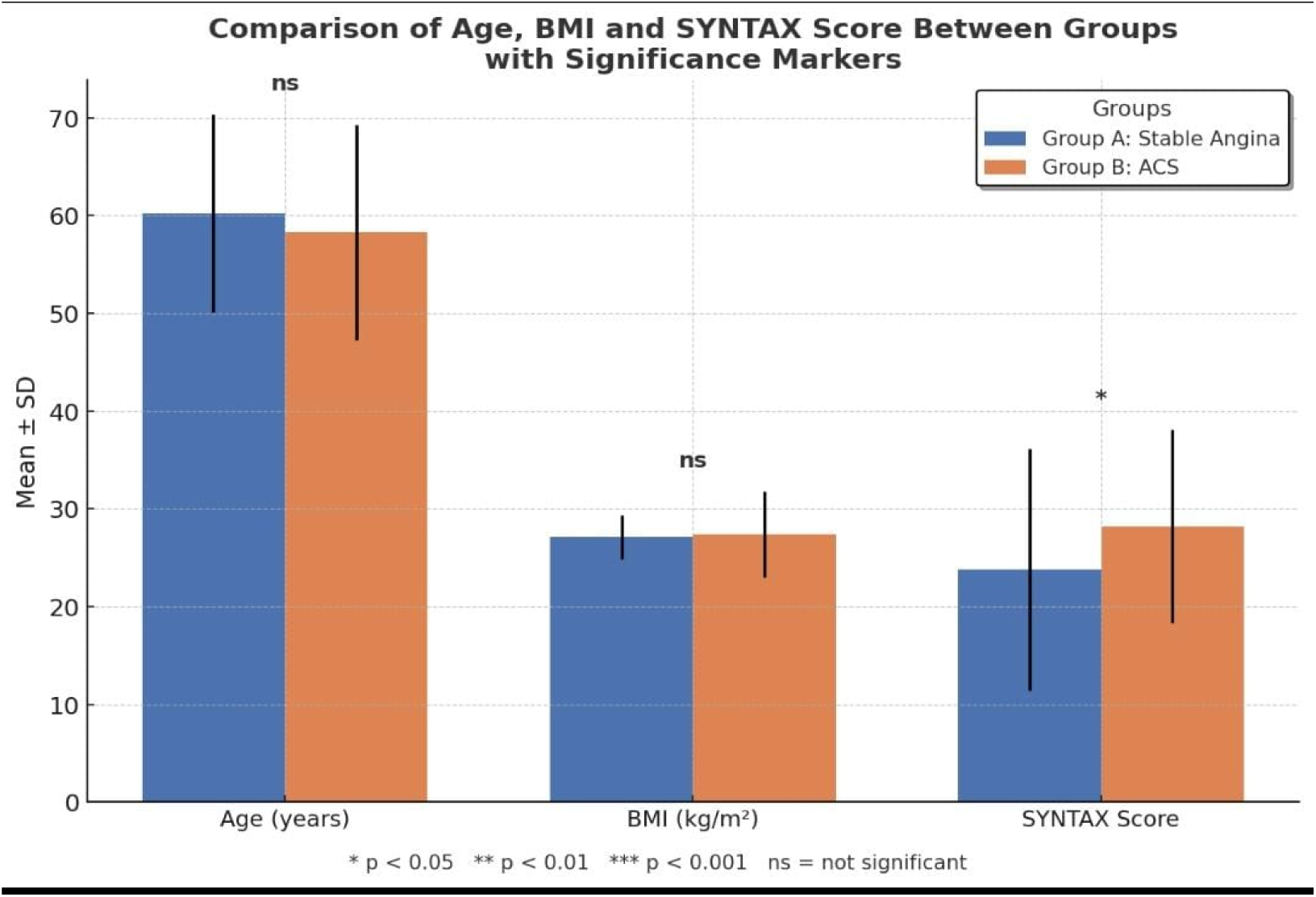
Comparison of mean age, BMI, and SYNTAX score (mean ± SD) between Group A (Stable Angina) and Group B (Acute Coronary Syndrome, ACS). No significant differences were observed in age and BMI between groups (ns = not significant). However, the mean SYNTAX score was significantly higher in the ACS group compared to the stable angina group (*p < 0.05).* Error bars indicate standard deviation

## DISCUSSION

In this study, we have compared the angiographic findings in patients reported with stable angina and acute coronary syndrome by using the SYNTAX score. The objective of this study is to compare the angiographic findings in stable angina with acute coronary syndrome by using SYNTAX score. Our results showed that the SYNTAX score was significantly higher in acute coronary syndrome patients as compared to stable angina patients. Recent advances in technology have led to an increase in the proportion of patients with ACS undergoing percutaneous coronary intervention (PCI). The selection of the optimal revascularization modality for patients with ACS depends on the patient characteristics and lesion severity. In addition, coronary artery bypass graft is still appropriate for patients with disease refractory to medical therapy and when PCI is not suitable or possible (Godoy, Lucas C., et al., 2019).

The complex coronary artery anatomy may be quantified in recognition of the SYNTAX score, which is solely generated from the coronary anatomy and lesion characteristics (Vizirianakis, Ioannis S., et al., 2021). The score, which was a crucial component of the SYNTAX trial design (Thuijs, Daniel JFM, et al., 2019), was initially created as a way to guarantee that the cardiologist and cardiac surgeon accurately reviewed the angiogram of patients with complex CAD, allowing a consensus to be reached regarding the best procedure and level of revascularization. It’s significant that the SYNTAX score was determined a priori, before the results of revascularization were known. In order to help with revascularization decisions, the SYNTAX trial results showed that the score is crucial in stratifying patients with complicated CAD (Gallo, Michele, et al., 2022).

The SYNTAX score of zero implies a low risk of cardiovascular, and generally promotes a more conservative approach to care, according to a study done by Berman et al. On the other hand, if the syntactic score is higher than 27, the risk is comparable to that of patients with clinically severe CAD, and aggressive medical therapy is typically needed (Berman, Daniel S et al., 2016).

Long-term outcome comparisons between patients with ACS and SAP among those receiving percutaneous coronary intervention (PCI) have produced mixed outcomes. The long-term risks of UAP and SAP following PCI were comparable in individuals with ACS and SAP in the Lescol Intervention Prevention Study (Sandeelo, Imran Khan, et al., 2019). The risks of major adverse cardiac events at 1 year between individuals with ACS and SAP were not significantly different, according to a sub analysis of the Arterial Revascularization Therapies Study (Matsoukis, Ioannis L., et al., 2021). However, compared to SAP, patients with ACS had a higher probability of death and combined death and myocardial infarction, according to Naidu and colleagues. Consideration should be given to the role of the overall atherosclerotic burden and disease progression following PCI in arteries not receiving intervention in ACS patients (Van Nguyen, Huy, et al., 2021). In the present study, a SYNTAX score was used to bifurcate the risk of adverse outcome in ACS versus stable angina patients. There was significantly high SYNTAX score was reported in ACS versus stable angina patients. Therefore, the risk of adverse outcome is significantly higher in ACS versus stable angina patients. Also, a high proportion of comorbidities were reported in patients in the ACS group as compared to stable angina patients. These results were comparable with the aforementioned studies. A few studies have assessed the predictive value SYNTAX score for long-term clinical outcomes in a PCI population (He, Jiqiang, et al., 2019; Cavalcante, Rafael, et al., 2017).

In addition, in this study, we have explored the SYNTAX score difference in patients with diabetes and hypertension. Furthermore, the syntax score was statistically significantly (p-value 0.001) high in patients with diabetes (31.4 ± 7.92) as compared to patients without diabetes (20.8 ± 11.9). Moreover, the SYNTAX score was significantly high in patients with hypertension (28.9 ± 9.05) as compared to patients without hypertension (17.2 ± 13.5). These results were comparable with the published data (Xu, Weifeng, et al., 2019; Kurtul BE, et al., 2021). In addition, another study conducted by (Kose, N., et al., 2019) reported a high mean difference of SYNTAX score in patients with stable angina versus ACS. Also, (Kose, N., et al., 2019) reported the high mean of SYNTAX score in patients with diabetes and hypertension (Kose, N., et al., 2019). These results were also comparable to our results. Furthermore, another study reported that a strong positive correlation exits between SYNTAX score and coronary artery disease; this means that the chance of severity of disease is higher with an increase in SYNTAX score. These results were also comparable to our data.

The SYNTAX score is unaffected by a patient’s clinical traits, some of which, such as patient age, have repeatedly been demonstrated to be independent predictors of mortality. In comparison to lesion-based scores like the American Heart Association lesion classification, prior research has shown that clinically based risk models, such as the Mayo Clinic Risk Score MCRS, perform better at predicting morbidity and mortality. A potential drawback to the SYNTAX Score’s application in risk stratification is the lack of any consideration for clinical factors in its calculation. Secondly, it was a single center study. Sample size was limited as we selected patients who were admitted to our department with cardiac issues and not all the patients admitted/visiting the hospital.

## CONCLUSION

SYNTAX score can be used to predict the complexity of CAD in patients. Therefore, all patients with low-to-intermediate pretest probability of CAD undergoing cardiac CT scan for disease assessment should have SYNTAX score calculated, and those with a greater SYNTAX score in the presence of significant disease are likely to have a high SYNTAX score on coronary angiography. The SYNTAX score helps a lot to the cardiologist and gives the proper result. It is very helpful to check the prognosis of the disease.

## Data Availability

Data Available uopn request

## Acknowledgement

Shaikh Zayed Post Graduate Medical Complex, for making this study easy to conduct.

## Financial Support

No financial support was needed.

## Declaration of Competing Interest

The authors declare no conflict of interest.

## Authors Contribution

S.A. and A.M. conceived the idea, and prepared the initial manuscript. A.S., S.A., R.K and M.A. collected the patient data and wrote the subsequent manuscript. M.A.I., S.A, M.H. refined and finalized the manuscript. Q.S. supervised the entire manuscript.

## Notes

### Competing Interest Statement

The authors have declared no competing interest.

### Funding Statement

self funded

### Author Declarations

Approval was taken from the Institutional Review Board (IRB) of Shaikh Zayed Medical Complex, Lahore

### Summary of Updates

The Number and sequence of authors are revised. The corresponding author has changed.

## REFERENCES

Al-Hadi, H. A., & Fox, K. A. (2009). Cardiac Markers in the Early Diagnosis and Management of Patients with Acute Coronary Syndrome. Sultan Qaboos University Medical Journal, 9(3), 231–246.

Abdelnabi M, Almaghraby A, Tok ÖÖ, Öz TK, Saleh Y, Morsi A, et al., A real-life correlation between clinical SYNTAX score II and carotid intima-media thickness in patients with stable coronary artery disease. J. Saudi Heart Assoc.. 2020;32(1):8.

Altekin, R. E., Kilinc, A. Y., Onac, M., & Cicekcibasi, O. (2020). Prognostic Value of the Residual SYNTAX Score on In-Hospital and Follow-Up Clinical Outcomes in ST Elevation Myocardial Infarction Patients Undergoing Percutaneous Coronary Interventions. Cardiology Research and Practice, 2020, 9245431. 10.1155/2020/9245431

Anstey Watkins, J., Wagner, F., Xavier Gómez-Olivé, F., Wertheim, H., Sankoh, O., & Kinsman, J. (2019). Rural South African Community Perceptions of Antibiotic Access and Use: Qualitative Evidence from a Health and Demographic Surveillance System Site. The American Journal of Tropical Medicine and Hygiene, 100(6), 1378–1390. 10.4269/ajtmh.18-0171

Banks, K., Lo, M., & Khera, A. (2010). Angina in Women without Obstructive Coronary Artery Disease. Current Cardiology Reviews, 6(1), 71–81. 10.2174/157340310790231608

Basit, H., Malik, A., & Huecker, M. R. (2022). Non ST Segment Elevation Myocardial Infarction. In StatPearls. StatPearls Publishing. http://www.ncbi.nlm.nih.gov/books/NBK513228/

Bergmark, B. A., Mathenge, N., Merlini, P. A., Lawrence-Wright, M. B., & Giugliano, R. P. (2022). Acute coronary syndromes. *Lancet (London*, England), 399(10332), 1347–1358. 10.1016/S0140-6736(21)02391-6

Boyette, L. C., & Manna, B. (2022). Physiology, Myocardial Oxygen Demand. In StatPearls. StatPearls Publishing. http://www.ncbi.nlm.nih.gov/books/NBK499897/

Berman DS, Arnson Y, Rozanski A. Coronary artery calcium scanning: the Agatston score and beyond. JACC: Circ. Cardiovasc. Imaging. 2016 Dec;9(12):1417–9.

Brilakis, E. (2021). Chapter 6—Coronary angiography. In E. Brilakis (Ed.), Manual of Percutaneous Coronary Interventions (pp. 97–109). Academic Press. 10.1016/B978-0-12-819367-9.00006-8

Budoff, M. J., Lakshmanan, S., Toth, P. P., Hecht, H. S., Shaw, L. J., Maron, D. J., Michos, E. D., Williams, K. A., Nasir, K., Choi, A. D., Chinnaiyan, K., Min, J., & Blaha, M. (2022). Cardiac CT angiography in current practice: An American society for preventive cardiology clinical practice statementZI. American Journal of Preventive Cardiology, 9, 100318. 10.1016/j.ajpc.2022.100318

Cassar, A., Holmes, D. R., Rihal, C. S., & Gersh, B. J. (2009). Chronic Coronary Artery Disease: Diagnosis and Management. Mayo Clinic Proceedings, 84(12), 1130–1146. 10.4065/mcp.2009.0391

Cirakoglu, O. F., Aslan, A. O., Akyuz, A. R., Kul, S., Şahin, S., Korkmaz, L., & Sayın, M. R. (2019). The value of syntax score to predict newLonset atrial fibrillation in patients with acute coronary syndrome. Annals of Noninvasive Electrocardiologyl: The Official Journal of the International Society for Holter and Noninvasive Electrocardiology, Inc, 24(4), e12622. 10.1111/anec.12622

Cavalcante R, Sotomi Y, Mancone M, Whan Lee C, Ahn JM, Onuma Y, et al., Impact of the SYNTAX scores I and II in patients with diabetes and multivessel coronary disease: a pooled analysis of patient level data from the SYNTAX, PRECOMBAT, and BEST trials. European heart journal. 2017 Jul 1;38(25):1969–77.

Costopoulos, C., Huang, Y., Brown, A. J., Calvert, P. A., Hoole, S. P., West, N. E. J., Gillard, J. H., Teng, Z., & Bennett, M. R. (2017). Plaque Rupture in Coronary Atherosclerosis Is Associated With Increased Plaque Structural Stress. Jacc. Cardiovascular Imaging, 10(12), 1472–1483. 10.1016/j.jcmg.2017.04.017

Eickhoff, M., Schüpke, S., Khandoga, A., Fabian, J., Baquet, M., Jochheim, D., Grundmann, D., Thienel, M., Bauer, A., Theiss, H., Brunner, S., Hausleiter, J., Massberg, S., & Mehilli, J. (2018). Age-dependent impact of the SYNTAX-score on longer-term mortality after percutaneous coronary intervention in an all-comer population. Journal of Geriatric Cardiologyl: JGC, 15(9), 559–566. 10.11909/j.issn.1671-5411.2018.09.009

Gillen, C., & Goyal, A. (2022). Stable Angina. In StatPearls. StatPearls Publishing. http://www.ncbi.nlm.nih.gov/books/NBK559016/

Godoy LC, Lawler PR, Farkouh ME, Hersen B, Nicolau JC, Rao V. Urgent revascularization strategies in patients with diabetes mellitus and acute coronary syndrome. CJC. 2019 Aug 1;35(8):993–1001.

Gallo M, Blitzer D, Laforgia PL, Doulamis IP, Perrin N, Bortolussi G, et al., Percutaneous coronary intervention versus coronary artery bypass graft for left main coronary artery disease: a meta-analysis. J. Thorac. Cardiovasc. 2022 Jan 1;163(1):94–105.

Ginghina, C., Ungureanu, C., Vladaia, A., Popescu, B. A., & Jurcut, R. (2009). The electrocardiographic profile of patients with angina pectoris. Journal of Medicine and Life, 2(1), 80–91.

He, Y.-M., Shen, L., & Ge, J.-B. (2020). Fallacies and Possible Remedies of the SYNTAX Score. Journal of Interventional Cardiology, 2020, 8822308. 10.1155/2020/8822308

He J, Zhao H, Yu X, Li Q, Lv S, Chen F, et al., SYNTAX score predicts long-term mortality in patients who underwent left main percutaneous coronary intervention treated with second-generation drug-eluting stents. International heart journal. 2017;58(3):344–50.

Hickam, D. H. (1990). Chest Pain or Discomfort. In H. K. Walker, W. D. Hall, & J. W. Hurst (Eds.), Clinical Methods: The History, Physical, and Laboratory Examinations (3rd ed.). Butterworths. http://www.ncbi.nlm.nih.gov/books/NBK416/

Kim, C. (2019). Risk Stratification by SYNTAX Score Systems in Current Percutaneous Revascularization Era. Korean Circulation Journal, 50(1), 35–37. 10.4070/kcj.2019.0348

Kim, Y.-H., Park, D.-W., Kim, W.-J., Lee, J.-Y., Yun, S.-C., Kang, S.-J., Lee, S.-W., Lee, C. W., Park, S.-W., & Park, S.-J. (2010). Validation of SYNTAX (Synergy between PCI with Taxus and Cardiac Surgery) score for prediction of outcomes after unprotected left main coronary revascularization. JACC. Cardiovascular Interventions, 3(6), 612–623. 10.1016/j.jcin.2010.04.004

Kirresh, A., White, L., Mitchell, A., Ahmad, S., Obika, B., Davis, S., Ahmad, M., & Candilio, L. (2022). Radiation-induced coronary artery disease: A difficult clinical conundrum. Clinical Medicine, 22(3), 251–256. 10.7861/clinmed.2021-0600

Kristin Newby, L. (2010). Chapter 25—Acute Coronary Syndromes. In G. S. Ginsburg & H. F. Willard (Eds.), Essentials of Genomic and Personalized Medicine (pp. 303–312). Academic Press. 10.1016/B978-0-12-374934-5.00025-8

Kurtul BE, Kurtul A, Yalçın F. Predictive value of the SYNTAX score for diabetic retinopathy in stable coronary artery disease patients with a concomitant type 2 diabetes mellitus. J. Diabetes Res. 2021 Jul 1;177:108875.

Kose N, Akin F, Yildirim T, Ergun G, Altun I. The association between the lymphocyte-to-monocyte ratio and coronary artery disease severity in patients with stable coronary artery disease. Eur Rev Med Pharmacol Sci. 2019 Mar 1;23(6):2570–5.

Kumar, A., & Cannon, C. P. (2009). Acute Coronary Syndromes: Diagnosis and Management, Part I. Mayo Clinic Proceedings, 84(10), 917–938.

Layne, K., & Ferro, A. (2017). Antiplatelet Therapy in Acute Coronary Syndrome. European Cardiology Review, 12(1), 33–37. 10.15420/ecr.2016:34:2

Lee, K. Y., Hwang, B.-H., Kim, C. J., Sa, Y. K., Choi, Y., Kim, J.-J., Choo, E.-H., Lim, S., Choi, I. J., Park, M.-W., Oh, G. C., Yang, I.-H., Yoo, K. D., Chung, W. S., & Chang, K. (2022). Prognostic Impact of the HFA-PEFF Score in Patients with Acute Myocardial Infarction and an Intermediate to High HFA-PEFF Score. Journal of Clinical Medicine, 11(15), 4589. 10.3390/jcm11154589

Li, X.-Q., Yin, C., Li, X.-L., Wu, W.-L., & Cui, K. (2021). Comparison of the prognostic value of SYNTAX score and clinical SYNTAX score on outcomes of Chinese patients underwent percutaneous coronary intervention. BMC Cardiovascular Disorders, 21, 334. 10.1186/s12872-021-02144-w

Manda, Y. R., & Baradhi, K. M. (2022). Cardiac Catheterization Risks and Complications. In StatPearls. StatPearls Publishing. http://www.ncbi.nlm.nih.gov/books/NBK531461/

Matsoukis IL, Karanasos A, Patsa C, AnousakisLVlachochristou N, Triantafyllou K, Kantzanou M. et al., LongLterm clinical outcomes of coronary artery bypass graft surgery compared to those of percutaneous coronary intervention with second generation drug eluting stents in patients with stable angina and an isolated lesion in the proximal left anterior descending artery. Catheter Cardiovasc Interv. 2021 Sep;98(3):447–57.

Mo, Y., & Xing, B. (2021). Correlation between coronary CTA-SYNTAX score and invasive coronary angiography-SYNTAX score. Zhong Nan Da Xue Xue Bao. Yi Xue Ban = Journal of Central South University. Medical Sciences, 46(8), 884–888. 10.11817/j.issn.1672-7347.2021.200837

null, null, Braunwald, E., Antman, E. M., Beasley, J. W., Califf, R. M., Cheitlin, M. D., Hochman, J. S., Jones, R. H., Kereiakes, D., Kupersmith, J., Levin, T. N., Pepine, C. J., Schaeffer, J. W., Smith, E. E., Steward, D. E., Theroux, P., Gibbons, R. J., Alpert, J. S., Eagle, K. A., … Smith, S. C. (2000). ACC/AHA Guidelines for the Management of Patients With Unstable Angina and Non–ST-Segment Elevation Myocardial Infarction: Executive Summary and Recommendations. Circulation, 102(10), 1193–1209. 10.1161/01.CIR.102.10.1193

Pakkal, M., Raj, V., & Mccann, G. P. (2011). Non-invasive imaging in coronary artery disease including anatomical and functional evaluation of ischaemia and viability assessment. The British Journal of Radiology, 84(Spec Iss 3), S280–S295. 10.1259/bjr/50903757

Parikh, R., Patel, A., Lu, B., Senapati, A., Mahmarian, J., & Chang, S. M. (2020). Cardiac Computed Tomography for Comprehensive Coronary Assessment: Beyond Diagnosis of Anatomic Stenosis. Methodist DeBakey Cardiovascular Journal, 16(2), 77–85. 10.14797/mdcj-16-2-77

Ramjattan, N. A., Lala, V., Kousa, O., & Makaryus, A. N. (2022). Coronary CT Angiography. In StatPearls. StatPearls Publishing. http://www.ncbi.nlm.nih.gov/books/NBK470279/

Rehman, I., Kerndt, C. C., & Rehman, A. (2022). Anatomy, Thorax, Heart Left Anterior Descending (LAD) Artery. In StatPearls. StatPearls Publishing. http://www.ncbi.nlm.nih.gov/books/NBK482375/

Safarian, H., Alidoosti, M., Shafiee, A., Salarifar, M., Poorhosseini, H., & Nematipour, E. (2014a). The SYNTAX Score Can Predict Major Adverse Cardiac Events Following Percutaneous Coronary Intervention. Heart Viewsl: The Official Journal of the Gulf Heart Association, 15(4), 99–105. 10.4103/1995-705X.151081

Sandeelo IK, Ahmed F, Kakepoto N, Shah MA, Parkash C, Ali H. To Assess the Beneficial Effects of Rosuvastatin Before Percutaneous Coronary Intervention in Patients with Acute Coronary Syndrome. Nat. Edi. Advi. Brd. 2019 Oct;30(10):60–4.

Safarian, H., Alidoosti, M., Shafiee, A., Salarifar, M., Poorhosseini, H., & Nematipour, E. (2014b). The SYNTAX Score Can Predict Major Adverse Cardiac Events Following Percutaneous Coronary Intervention. Heart Views: The Official Journal of the Gulf Heart Association, 15(4), 99–105. 10.4103/1995-705X.151081

Salvatore, A., Boukhris, M., Giubilato, S., Tomasello, S. D., Castaing, M., Giunta, R., Marzà, F., Abdelbasset, H. M., Khamis, H., & Galassi, A. R. (2016). Usefulness of SYNTAX score II in complex percutaneous coronary interventions in the setting of acute coronary syndrome. Journal of the Saudi Heart Association, 28(2), 63–72. 10.1016/j.jsha.2015.07.003

Sarkees, M. L., & Bavry, A. A. (2009). Acute coronary syndrome (unstable angina and non-ST elevation MI). BMJ Clinical Evidence, 2009, 0209.

Scherff, F., Vassalli, G., Sürder, D., Mantovani, A., Corbacelli, C., Pasotti, E., Klersy, C., Auricchio, A., Moccetti, T., & Pedrazzini, G. B. (2011). The SYNTAX score predicts early mortality risk in the elderly with acute coronary syndrome having primary PCI. The Journal of Invasive Cardiology, 23(12), 505–510.

Serruys, P. W., Onuma, Y., Garg, S., Sarno, G., van den Brand, M., Kappetein, A.-P., Van Dyck, N., Mack, M., Holmes, D., Feldman, T., Morice, M.-C., Colombo, A., Bass, E., Leadley, K., Dawkins, K. D., van Es, G.-A., Morel, M.-A. M., & Mohr, F. W. (2009). Assessment of the SYNTAX score in the Syntax study. EuroIntervention: Journal of EuroPCR in Collaboration with the Working Group on Interventional Cardiology of the European Society of Cardiology, 5(1), 50–56. 10.4244/eijv5i1a9

Shahjehan, R. D., & Bhutta, B. S. (2022). Coronary Artery Disease. In StatPearls. StatPearls Publishing. http://www.ncbi.nlm.nih.gov/books/NBK564304/

Shalev, A., Nakazato, R., Arsanjani, R., Nakanishi, R., Park, H.-B., Otaki, Y., Cheng, V. Y., Gransar, H., LaBounty, T. M., Hayes, S. W., Berman, D. S., & Min, J. K. (2016). SYNTAX Score Derived From Coronary CT Angiography for Prediction of Complex Percutaneous Coronary Interventions. Academic Radiology, 23(11), 1384–1392. 10.1016/j.acra.2016.07.003

Suh, Y. J., Han, K., Chang, S., Kim, J. Y., Im, D. J., Hong, Y. J., Lee, H.-J., Hur, J., Kim, Y. J., & Choi, B. W. (2017). SYNTAX score based on coronary computed tomography angiography may have a prognostic value in patients with complex coronary artery disease. Medicine, 96(37), e7999. 10.1097/MD.0000000000007999

Sukhija, R., Aronow, W. S., Kakar, P., Garza, L., Sachdeva, R., Sinha, A., & Mehta, J. L. (2006). Relation of microalbuminuria and coronary artery disease in patients with and without diabetes mellitus. The American Journal of Cardiology, 98(3), 279–281.

Takagi, K., Tanaka, A., Yoshioka, N., Morita, Y., Yoshida, R., Kanzaki, Y., Watanabe, N., Yamauchi, R., Komeyama, S., Sugiyama, H., Shimojo, K., Imaoka, T., Sakamoto, G., Ohi, T., Goto, H., Ishii, H., Morishima, I., & Murohara, T. (2021). In-hospital mortality among consecutive patients with ST-Elevation myocardial infarction in modern primary percutaneous intervention era ∼ Insights from 15-year data of single-center hospital-based registry ∼. PLoS ONE, 16(6), e0252503. 10.1371/journal.pone.0252503

Thuijs DJ, Kappetein AP, Serruys PW, Mohr FW, Morice MC, Mack MJ, et al. Percutaneous coronary intervention versus coronary artery bypass grafting in patients with, three-vessel or left main coronary artery disease: 10-year follow-up of the multicentre randomised controlled SYNTAX trial. The Lancet. 2019 Oct 12;394(10206):1325–34.

Villa, A. D., Sammut, E., Nair, A., Rajani, R., Bonamini, R., & Chiribiri, A. (2016). Coronary artery anomalies overview: The normal and the abnormal. World Journal of Radiology, 8(6), 537–555. 10.4329/wjr.v8.i6.537

Van Nguyen H, Khuong LQ, Nguyen AT, Nguyen AL, Nguyen CT, Nguyen HT, et al., Changes in, and predictors of, quality of life among patients with unstable angina after percutaneous coronary intervention. J Eval Clin Pract. 2021 Apr;27(2):325–32.

Vizirianakis IS, Chatzopoulou F, Papazoglou AS, Karagiannidis E, Sofidis G, Stalikas N, et al., The GEnetic Syntax Score: a genetic risk assessment implementation tool grading the complexity of coronary artery disease—rationale and design of the GESS study. BMC Cardiovascular Disorders. 2021 Dec;21(1):1–9.

Xu W, Guan H, Gao D, Pan J, Wang Z, Alam M, et al. Sex-specific association of monocyte count to high-density lipoprotein ratio with SYNTAX score in patients with suspected stable coronary artery disease. Medicine. 2019 Oct;98(41).

Yang, C., Deng, Z., Li, J., Ren, Z., & Liu, F. (2021). Meta-analysis of the relationship between interleukin-6 levels and the prognosis and severity of acute coronary syndrome. Clinics, 76, e2690. 10.6061/clinics/2021/e2690

Yüceler, Z., Kantarcı, M., Tanboğa, İ. H., Sade, R., Kızrak, Y., Pirimoğlu, B., Bayraktutan, Ü., Oğul, H., & Aksakal, E. (2016). Coronary lesion complexity assessed by SYNTAX score in 256-slice dual-source MDCT angiography. Diagnostic and Interventional Radiology, 22(4), 334–340. 10.5152/dir.2015.15298

Zhang, Y., Iqbal, J., Campos, C., Klaveren, D., Bourantas, C., Dawkins, K., Banning, A., Escaned, J., Vries, T., Morel, M.-A., Farooq, V., Onuma, Y., Garcia-Garcia, H., Stone, G., Steyerberg, E., Mohr, F., & Serruys, P. (2014). Prognostic Value of Site SYNTAX Score and Rationale for Combining Anatomic and Clinical Factors in Decision Making. Journal of the American College of Cardiology, 64, 423–432. 10.1016/j.jacc.2014.05.022

